# α-synuclein seed amplification assay positivity is associated with motor progression in the prodromal stage independent of phenoconversion

**DOI:** 10.64898/2026.11.16.26368235

**Authors:** Xiao Deng, Mahsa Zeynali, Shermyn Neo Xiumin, Prakash Kumar Manharlal, Jing Han Ng, Seyed Ehsan Saffari, Louis CS Tan, Eng-King Tan, Bin Xiao

**Author notes:** **Correspondence**, Eng-King Tan, Department of Neurology, National Neuroscience Institute, Singapore., Bin Xiao, National Neuroscience Institute, Singapore. Xiao Deng & Mahsa Zeynali contributed equally to this study.

## Abstract

**Background:** α-Synuclein seed amplification assay (αSyn-SAA) has emerged as a promising biomarker for detecting underlying α-synuclein pathology and predicting phenoconversion in prodromal Parkinson’s disease (PD). However, its association with longitudinal clinical progression during the prodromal stage remains unclear. We investigated whether baseline αSyn-SAA status and kinetic parameters were associated with motor and cognitive progression and subsequent phenoconversion in prodromal individuals.

**Methods:** Clinical progression was assessed using the Movement Disorder Society-Sponsored Revision of the Unified Parkinson’s Disease Rating Scale (MDS-UPDRS) Part III motor scores, Hoehn and Yahr (H&Y) stage, and Montreal Cognitive Assessment (MoCA) scores. Linear mixed-effects models were utilized to examine the associations between baseline αSyn-SAA status, kinetic parameters, and longitudinal changes in these clinical measures. Additionally, time-to-event analyses were performed to assess the associations between SAA measurements and phenoconversion to PD or dementia with Lewy bodies.

**Results:** We identified 2,532 participants with available baseline αSyn-SAA status in the Parkinson’s Progression Markers Initiative (PPMI) prodromal cohort. This longitudinal study, which included up to 12 years of follow-up, comprised 1,714 αSyn-SAA-positive and 818 αSyn-SAA-negative individuals. Over the follow-up period, αSyn-SAA-positive participants exhibited significantly faster motor progression measured by MDS-UPDRS Part III motor score than αSyn-SAA-negative participants (0.611 vs. 0.139 points/year; P<0.001). This association persisted after excluding phenoconverters (0.430 vs. 0.202 points/year; P=0.014) and restricting follow-up to seven years (0.588 vs. 0.137 points/year; P<0.001). Among the 1,796 participants included in the time-to-event analyses, 181 phenoconversion events occurred. Baseline αSyn-SAA positivity was associated with a significantly higher hazard of phenoconversion (adjusted hazard ratio 5.72, 95% confidence interval 3.78-8.68; P<0.001), whereas kinetic parameters showed no significant associations with phenoconversion risk.

**Conclusions:** Baseline αSyn-SAA positivity was associated with accelerated motor progression during the prodromal stage independent of phenoconversion. These findings support the potential utility of αSyn-SAA positivity as a prognostic biomarker for motor progression and phenoconversion, with implications for risk stratification and participant selection in future disease-modifying trials.

## Introduction

Parkinson’s disease (PD) is pathologically characterized by Lewy bodies composed of misfolded α-synuclein and loss of dopaminergic neurons in the substantia nigra pars compacta (SNpc). Clinically, PD manifests with motor symptoms including resting tremor, bradykinesia, and rigidity, which form the basis of diagnostic criteria, as well as a broad spectrum of non-motor manifestations including cognitive impairment, autonomic dysfunction, sleep disturbances, and depression.^1^ Notably, many non-motor symptoms can emerge years before the onset of motor symptoms and have been recognized as features of the prodromal phase.^2,3^ Substantial nigrostriatal neurodegeneration has already occurred by the time of clinical diagnosis, with studies suggesting that over 50% of nigral dopaminergic neurons may be lost before the onset of overt motor symptoms.^4,5^ Thus, clinical early-stage PD may represent a relatively advanced pathological stage, by which point critical neurodegenerative thresholds may have been crossed and downstream pathogenic processes established.^6^ This may partially explain the limited clinical efficacy observed in trials of α-synuclein-targeted immunotherapies, including prasinezumab^7,8^ and cinpanemab,^9^ despite strong preclinical evidence and successful target engagement.^10^ The prodromal phase refers to a period during which underlying neurodegeneration has begun, but diagnostic criteria for a clinically manifest synucleinopathy have not yet been fulfilled. Because pathological changes may be less advanced and more amenable to intervention at this stage, the prodromal phase is increasingly considered a promising therapeutic window for disease-modifying interventions. Consequently, there is an unmet need for biomarkers that can improve risk stratification during the prodromal phase and inform the timely application of disease-modifying interventions.^11,12^

α-Synuclein seed amplification assay (αSyn-SAA) enables the detection of seeding-competent pathological α-synuclein in biological samples, including cerebrospinal fluid^13^ and skin tissues.^14^ The assay has emerged as a promising diagnostic marker for synucleinopathies, with sensitivities and specificities generally exceeding 85-90% in PD^15^. Studies have shown that αSyn-SAA positivity is detectable during the prodromal phase, including in individuals with isolated REM sleep behaviour disorder (iRBD),^16,17^ hyposmia^15,18^ and genetic risk factors for PD.^15^ Interestingly, αSyn-SAA positivity was lower in carriers of LRRK2 mutations.^15,19^ but higher in carriers of GBA mutations.^15,20^ This pattern mirrors the differing prevalence of Lewy body pathology reported in these genetic forms of PD, with Lewy body pathology being common in GBA-associated PD^21^ but less consistently observed in LRRK2-associated PD,^22^ which further supports the utility of αSyn-SAA as a biomarker of underlying α-synuclein pathology. In addition, αSyn-SAA positivity has been found to be associated with an increased likelihood of phenoconversion among prodromal participants,^17,23^ suggesting that αSyn-SAA possesses prognostic value during the prodromal stage. Given that αSyn-SAA directly detects seeding-competent pathological α-synuclein, which is a key driver of PD pathogenesis, αSyn-SAA is expected to predict disease progression. However, evidence linking αSyn-SAA to clinical progression remains inconsistent. While Orru et al. reported significant associations between αSyn-SAA kinetic measures and cognitive outcomes in PD,^24^ Schumacher et al. reported little association between αSyn-SAA parameters and clinical progression in PD, including motor and cognitive features.^25^

In essence, αSyn-SAA has established its role as a biomarker of underlying synuclein pathology, providing diagnostic value for synucleinopathies. αSyn-SAA may also predict phenoconversion risk in prodromal participants. However, whether αSyn-SAA serves as a prognostic biomarker of clinical progression in PD remains unclear. In particular, its association with clinical progression during the prodromal phase has not been investigated.

## Materials and Methods

### 1. Study population

Data used in the preparation of this article were obtained on July 29, 2026, from the Parkinson’s Progression Markers Initiative (PPMI) database (www.ppmi-info.org/access-data-specimens/download-data), RRID:SCR_006431. For up-to-date information on the study, visit www.ppmi-info.org. PPMI is an ongoing international, multicentre, observational cohort study that aims to identify clinical, imaging, and biological markers of Parkinson’s disease progression.^26^ The study is registered at ClinicalTrials.gov (NCT01141023; current PPMI Clinical protocol NCT04477785). Each participating site obtained approval from its local institutional review board or ethics committee, and all participants provided written informed consent. We recruited all participants from phase I and phase II of the Prodromal cohort. Details of the eligibility criteria for both phases are available on the PPMI website (https://www.ppmi-info.org).

### 2. Study variables pre-processing

This was a longitudinal cohort study with up to 12 years of follow-up. We extracted demographic, clinical, cerebrospinal fluid (CSF), and blood biomarker variables for analysis. Demographic variables included age, sex, years of education, race, family history of PD, and body mass index (BMI). Blood-based variables comprised the genetic factor apolipoprotein E ε4 (APOE ε4) status, alongside serum uric acid, serum glucose, and serum creatinine to account for underlying metabolic comorbidities such as gout, diabetes mellitus (DM), and kidney disease. Clinical variables encompassed Montreal Cognitive Assessment (MoCA) score, Hoehn and Yahr stage (H&Y), and Movement Disorder Society– Unified Parkinson’s Disease Rating Scale (MDS-UPDRS) Part III motor score.

Baseline CSF α-synuclein seed amplification assay (SAA) data were obtained from three assay projects: Project 237 (24-hour assay), Project 155 (150-hour assay), and Project 299 (35-hour assay). Both qualitative and quantitative SAA parameters were extracted to characterize α-synuclein aggregation properties. CSF α-synuclein SAA status was defined as a binary variable indicating the positivity or negativity of α -synuclein aggregates based on the newest available assay protocol, whereas inconclusive or missing/unknown results were excluded.

As kinetic values are protocol-dependent, kinetic measures were not pooled across assays. Three baseline SAA kinetic measures were extracted from Project 237, including maximum fluorescence (Fmax), time to threshold (TTT), and area under the fluorescence curve (AUC). Kinetic values from up to three technical replicates were summarized by their arithmetic mean, and when a baseline sample had been rerun, only the most recent run date was used. Kinetic parameters were further dichotomized accordingly to their median values among SAA-positive participants. Figure 1 outlines the overall data preparation and preprocessing framework.

**Figure 1.**
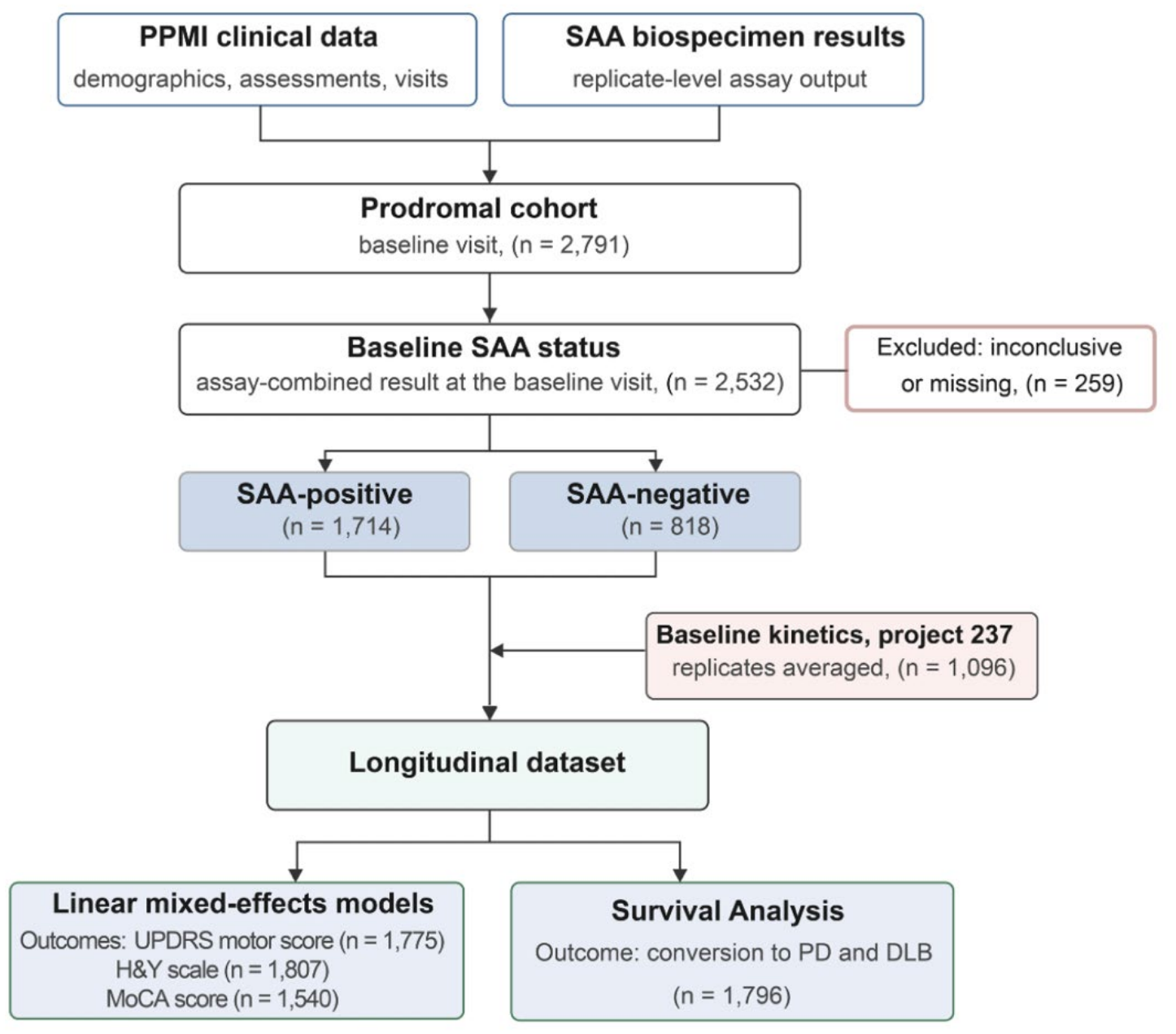
Study design and data pre-processing workflow and analytic plan.

### 3. Statistical analysis

Descriptive statistics were summarized as mean and standard deviation (SD) for continuous variables and as counts and percentages for categorical variables. Baseline differences between the SAA-positive and SAA-negative groups were assessed using the Mann–Whitney U test for continuous variables and the chi-square test or Fisher’s exact test for categorical variables, as appropriate based on expected cell counts.

#### Linear Mixed-Effects Models (Primary Analysis)

Longitudinal changes in clinical trajectories, specifically the MDS-UPDRS Part III motor score, H&Y stage, and MoCA score, over the 12-year follow-up period were analysed using linear mixed-effects models (LMMs). Time was modelled as a continuous variable (visit year) to estimate the annual rate of progression. The models included fixed effects for time, SAA groupings (SAA binary status or kinetic groups), and their interaction (group × time). To account for individual variability at baseline values and rates of change, participant-specific random intercepts and random slopes were included using an unstructured covariance matrix. All models were adjusted for potential confounders, including baseline age, sex, family history of PD, the baseline score of the outcome being measured, and incident conversion to a clinical diagnosis of PD or Dementia with Lewy bodies (DLB) during the follow-up period. For models specifically assessing cognitive progression (MoCA scores), years of education and APOE *ε*4status were included as additional covariates. Models were fitted using restricted maximum likelihood (REML) estimation. The assumption of normality was assessed visually using quantile-quantile (Q-Q) plots of the standardized residuals. The disease progression slopes, evaluated by the interaction term of group × time, were reported as adjusted beta coefficients with 95% confidence intervals (CIs).

To ensure the robustness of our primary longitudinal findings, two sensitivity analyses were performed. First, to eliminate the confounding effect that worsened motor trajectories were driven solely by incident PD or DLB, we excluded all participants who converted and compared the longitudinal trajectories strictly between SAA-positive and SAA-negative non-converters. Second, to account for potential attrition bias and missing data in the later years of the 12-year study period, we restricted the longitudinal follow-up duration to 7 years and re-estimated the models.

#### Survival Analysis (Secondary Analysis)

To complement the longitudinal trajectory data, time-to-event analyses were performed to evaluate the association of baseline SAA status (positive vs. negative) and kinetic parameters (Fmax, AUC, TTT) with subsequent phenoconversion to a site investigator’s primary diagnosis of PD or DLB. Follow-up time was calculated from baseline to the date of the first qualifying diagnosis. Participants without an event were censored at their last observed clinical visit. Individuals who already met the criteria for a qualifying diagnosis at baseline were excluded from the incident-event analyses. Phenoconversion-free survival was estimated using the Kaplan–Meier method, with group differences assessed via two-sided log-rank tests. The proportional hazards assumption was verified for all models using Schoenfeld residual-based tests. Cox proportional hazards regression models were utilized to calculate hazard ratios (HRs) and 95% CIs for the associations between SAA parameters and phenoconversion, adjusting for baseline age and sex.

The significance level was set at p < 0.05. Dataset processing and statistical analyses were conducted in Python (version 3.13.15) using the pandas (version 2.2.3), NumPy (version 2.1.3), SciPy (version 1.16.3), statsmodels (version 0.14.6), and lifelines (version 0.30.3) packages.

## Results

### Baseline characteristics

A total of 2532 participants from the prodromal cohort were included in the analysis, of whom 1714 (67.7%) were SAA-positive, and 818 (32.3%) were SAA-negative. Baseline demographic and clinical characteristics are summarized in Table 1. Compared with SAA-negative individuals, SAA-positive participants were significantly older, had a higher proportion of males, reported fewer years of education, and had a higher prevalence of APOE *ε*4 carriers and were less likely to have a family history of PD. Clinically, SAA-positive participants demonstrated greater baseline impairment, including significantly lower MoCA scores, higher Hoehn and Yahr stages, and higher MDS-UPDRS Part III motor scores. There were no significant differences between the groups regarding race, BMI, serum hyperuricemia, elevated serum glucose, or elevated serum creatinine.

**Table 1.** Baseline characteristics between SAA-positive and SAA-negative groups in the prodromal cohort.

| **Characteristic** | **SAA-positive**<br>**(n = 1714)** | **SAA-negative**<br>**(n = 818)** | **P-value** |
|---|---|---|---|
| Age at baseline (years) | 68.51 ± 6.12 | 64.22 ± 6.99 | <0.001** |
| Male sex, (n, %) | 939 (54.8) | 310 (37.9) | <0.001** |
| Years of education | 16.31 ± 2.70 | 16.47 ± 2.83 | 0.039* |
| White race (n, %) | 1620/1698 (95.4) | 768/809 (94.9) | 0.673 |
| Family history of PD (n, %) | 587/1708 (34.4) | 513/815 (62.9) | <0.001** |
| APOE ε4 carriers (n, %) | 263/906 (29.0%) | 174/739 (23.5%) | 0.03* |
| BMI (kg/m²) | 27.60 ± 7.49 | 27.88 ± 5.75 | 0.241 |
| Hyperuricaemia (n, %) | 77/674 (11.4%) | 26/309 (8.4%) | 0.187 |
| Elevated serum glucose (n, %) | 48/676 (7.1%) | 32/311 (10.3%) | 0.114 |
| Elevated serum creatinine (n, %) | 12/674 (1.8%) | 6/308 (1.9%) | 1 |
| MoCA score | 26.60 ± 2.45 | 26.79 ± 2.40 | 0.03* |
| H&Y stage | 0.23 ± 0.59 | 0.13 ± 0.46 | <0.001** |
| MDS-UPDRS Part III motor score | 5.06 ± 6.06 | 3.57 ± 4.62 | <0.001** |

**Abbreviations:** SAA: seed amplification assay; APOE: apolipoprotein E ε4; BMI: Body mass index; Hyperuricemia: Serum uric acid level ≥404 μmol/L; Elevated serum glucose: Serum glucose level ≥7.0 mmol/L; Elevated serum creatinine: Serum creatinine level above sex-specific reference limits; MoCA: Montreal Cognitive Assessment; H&Y: Hoehn and Yahr; MDS-UPDRS: Movement Disorder Society–Unified Parkinson’s Disease Rating Scale
\*: P- value <0.05; \*\*: P- value <0.001

### SAA positivity and the longitudinal clinical trajectories

In the primary analysis, over up to 12 years of longitudinal follow-up, the SAA-positive group demonstrated a significantly steeper annual increase in the MDS-UPDRS Part III motor score compared with the SAA-negative group (+0.611 vs. +0.139 points/year; P<0.001), after adjusting for age, sex, family history, phenoconversion status, and baseline MDS-UPDRS Part III score (Table 2, Figure 2A). The differences in the annual rates of change in H&Y stage (P=0.164) and MoCA scores (P=0.924) were not significant between the two groups.

**Table 2.** Comparison of motor progression rates between SAA-positive and negative groups.

| MDS-UPDRS Part III | group | Slope estimation | 95% CI |  | P-value† |
|---|---|---|---|---|---|
|  |  |  | Lower | Upper |  |
| Main analyses | SAA negative (ref) | 0.139 | 0.008 | 0.271 | <0.001** |
|  | SAA positive | 0.611 | 0.381 | 0.841 |  |
| Sensitivity analyses 1 | SAA negative (ref) | 0.202 | 0.123 | 0.282 | 0.014 * |
|  | SAA positive | 0.430 | 0.265 | 0.595 |  |
| Sensitivity analyses 2 | SAA negative (ref) | 0.137 | 0.005 | 0.269 | <0.001** |
|  | SAA positive | 0.588 | 0.366 | 0.809 |  |

Abbreviations: SAA: seed amplification assay; MDS-UPDRS: Movement Disorder Society–Unified Parkinson’s Disease Rating Scale.
† Linear mixed-effects models were used to compare longitudinal changes in UPDRS Part III motor scores between SAA-positive and SAA-negative participants.
\*: P- value <0.05; \*\*: P- value <0.001

**Figure 2.**
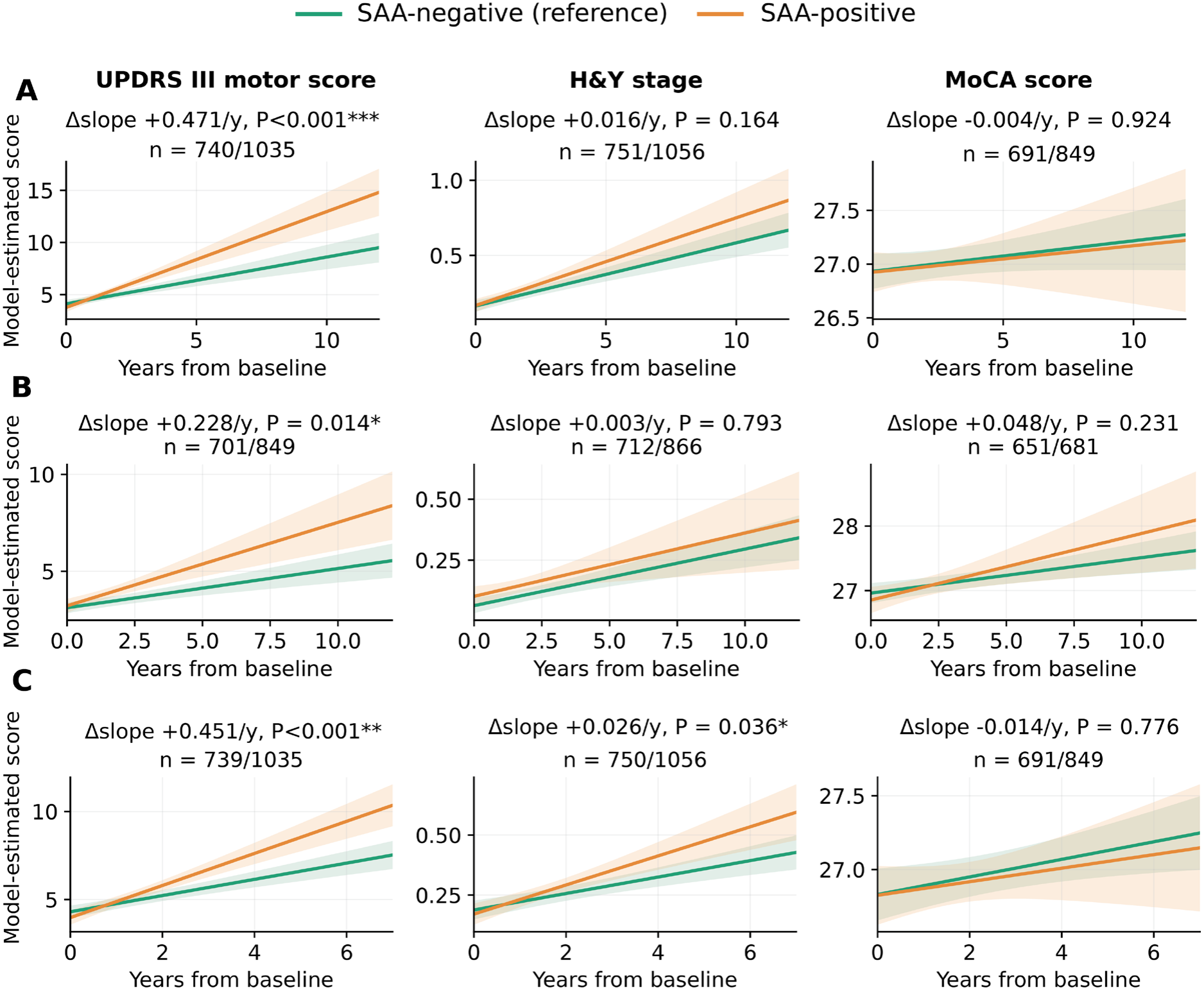
Comparison of longitudinal clinical progression rates between SAA-positive and SAA-negative groups. Model-estimated trajectories for MDS-UPDRS Part III motor score, Hoehn and Yahr stage, and MoCA score are shown for SAA-positive and SAA-negative participants. Δslope represents the estimated difference in the annual rate of change between SAA-positive and SAA-negative participants; P values test the difference in slopes.(A) Main analysis, adjusted for confounders including age, sex, family history, baseline assessment score, and phenoconversion status. (B) Sensitivity analysis 1 restricted to participants who did not phenoconvert during follow-up. (C) Sensitivity analysis 2 restricted to the first 7 years of follow-up, using the same covariates as in the main analysis.

Two sensitivity analyses were performed. In the first sensitivity analysis, which excluded phenoconverters, SAA-positive non-converters continued to show significantly faster progression in the MDS-UPDRS Part III motor score compared with SAA-negative non-converters (+0.430 vs. +0.202 points/year; P=0.014; Table 2, Figure 2B). In the second sensitivity analysis, follow-up was restricted to the first 7 years to minimize the impact of high attrition at later time points. These findings were consistent with the main analysis. The SAA-positive group maintained a significantly steeper progression rate in MDS-UPDRS Part III motor score compared with the SAA-negative group after adjustment for age, sex, family history, phenoconversion status, and baseline MDS-UPDRS Part III score (+0.588 vs. +0.137 points/year; P<0.001; Table 2, Figure 2C).

In terms of the association between SAA kinetic groupings (baseline Fmax, TTT, and AUC) and the longitudinal clinical trajectories of MDS-UPDRS Part III motor score, H&Y stage, and MoCA score, no significant differences in longitudinal slopes were observed between any SAA kinetic groups and clinical outcomes (all P > 0.05, Figure S1, Supplementary results1).

### SAA status, kinetic parameters, and phenoconversion

For our secondary analysis, the updated dataset featured a substantially larger prodromal cohort (1,796 vs. 96 participants), a higher number of phenoconversion events (181 vs. 23), and a longer follow-up period (12 vs. 9.2 years) compared to the study by Coughlin et al.^23^ A detailed comparison of the survival analyses before and after the PPMI cohort expansion is presented in (Table S1, Supplementary results 2).

During the 12-year follow-up of 1,796 participants, 181 phenoconversion events were recorded (148 among 1,040 SAA-positive and 33 among 756 SAA-negative individuals). Phenoconversion-free survival was significantly lower for SAA-positive participants (log-rank test, P<0.001; Figure 3A). After adjustment for baseline age and sex, SAA positivity was significantly associated with a higher hazard of phenoconversion (HR 5.72, 95% CI 3.78–8.68, P<0.001; Table S1, Figure 3B). Within the subset of 991 SAA-positive participants with kinetic measurements, the previously reported difference between slow and fast TTT (P=0.03) lost statistical significance (adjusted HR 0.73, 95% CI 0.51–1.03, P=0.074; Table S1). Similarly, median-split Fmax and AUC were not significantly associated with phenoconversion after adjustment for baseline age and sex (Figure 3B-E).

**Figure 3.**
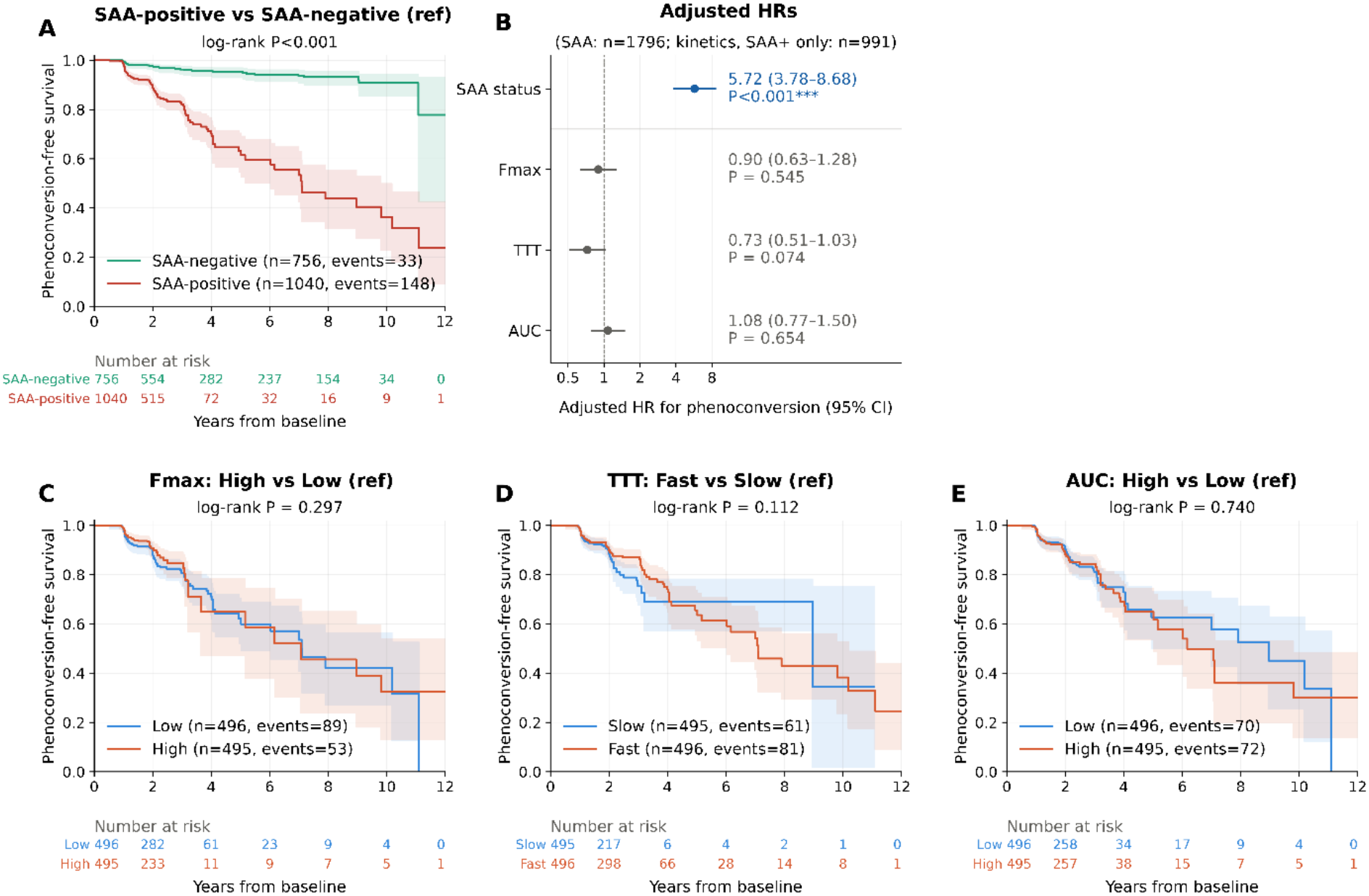
Phenoconversion-free survival and risk of phenoconversion according to baseline CSF SAA parameters. (A) Kaplan–Meier estimates of phenoconversion-free survival in the overall prodromal cohort according to baseline SAA binary status. (B) Forest plot showing adjusted HRs and 95% CIs for the associations of SAA status and kinetic parameters with phenoconversion risk, estimated using Cox proportional hazards models. (C–E) Kaplan–Meier estimates among SAA-positive participants with Project 237 kinetic data according to median-split Fmax (high vs low), TTT (fast vs slow), and AUC (high vs low). Shading indicates 95% CIs, and numbers at risk are shown below each Kaplan–Meier plot. P values for Kaplan–Meier analyses are from unadjusted log-rank tests.

## Discussion

To our knowledge, our findings provide the first evidence that αSyn-SAA has the potential to serve as a prognostic biomarker for clinical progression in the prodromal phase. Together with the development of αSyn-SAA as a potential diagnostic tool for PD,^27^ our study suggests that the assay has distinct significance across the disease stages. Utilizing updated PPMI data with a substantially expanded cohort (>2,500 participants) and up to 12 years of follow-up, we demonstrated that αSyn-SAA positivity is significantly associated with accelerated motor deterioration in prodromal individuals. This association remained robust in two sensitivity analyses, which excluded phenoconverters and restricted to shorter follow-up durations with less dropout, respectively.

In the primary analysis, our study found that αSyn-SAA positivity was associated with motor deterioration during the prodromal phase. By contrast, a previous study did not find a significant association between αSyn-SAA and motor progression in clinically diagnosed PD.^25^ This stage-dependent association of αSyn-SAA and clinical progression may provide insight into the evolving role of pathological α-synuclein in PD. One possible interpretation is that α-synuclein seeds play an important role in the earliest stage of disease when the pathogenic cascade is still developing. Accordingly, αSyn-SAA positivity may identify individuals at greater risk of motor progression during the prodromal phase. As disease advances, however, progression may become increasingly influenced by downstream pathogenic processes rather than α-synuclein seeding activity alone.^6,28^ Under this model, αSyn-SAA may reflect both pathology and disease progression during the prodromal stage, but primarily pathology in clinically manifest PD.

A major challenge when studying motor progression in prodromal subjects is the potential confounding introduced by phenoconverters, who experience sufficient motor worsening to cross the diagnostic threshold for PD. Consequently, observed association between αSyn-SAA positivity and motor progression could be driven primarily by phenoconverters. To address this concern, we performed sensitivity analyses excluding the phenoconverters. Although such analyses reduce sample size and may introduce selection bias, they represent a conservative strategy because they completely remove the most obvious source of confounding. Notably, the association between αSyn-SAA positivity and motor deterioration persisted within participants who remained prodromal throughout follow-up. Together with the primary analyses that adjusted for conversion status as a covariate, these findings support an association between α-synuclein seeding status and motor deterioration even among individuals who never cross the diagnostic threshold during the study period.

The present findings may have important implications for the development of disease-modifying therapies for PD. Our findings that αSyn-SAA positivity was associated with motor progression at the prodromal stage suggest that the role of pathological α-synuclein in PD pathogenesis may be greatest during this stage. Accordingly, α-synuclein-directed therapies may be most effective when administered at earlier stages of disease. The findings also support a role for αSyn-SAA in patient stratification. Because αSyn-SAA-positive prodromal individuals exhibit more rapid motor deterioration, disease progression in this population may be more closely linked to pathological α-synuclein. Consequently, these individuals may represent the subgroup most likely to benefit from α-synuclein-targeted interventions. As αSyn-SAA directly detects the pathological α-synuclein species targeted by many emerging therapies,^29,30^ the assay could serve as a biomarker for participant selection and cohort enrichment in future α-synuclein-targeted therapeutic trials. A further implication relates to clinical trial endpoints. Most trials involving prodromal participants have relied on phenoconversion as the primary outcome measure. However, phenoconversion requires prolonged follow-up and represents a dichotomous clinical milestone. Our findings suggest that quantitative measures of motor progression, such as MDS-UPDRS Part III scores, may provide sensitive and clinically meaningful indicators of treatment effect in αSyn-SAA-positive prodromal individuals. Thus, slowing motor deterioration may represent a meaningful therapeutic benefit in prodromal participants, even if phenoconversion is not significantly prevented during the trial period.

In our secondary analysis, we confirmed that α Syn-SAA positivity is associated with a higher likelihood of phenoconversion, reinforcing its value as a robust prognostic marker.^17, 23^ However, in our substantially expanded cohort (n=1,796, an approximate 20-fold increase from the original prodromal cohort of n= 96), we could not replicate the previously reported significant associations between TTT and phenoconversion.^23^ This discrepancy may arise from several methodological and biological factors. First, dichotomizing a continuous variable like TTT inherently discards nuanced data and diminishes statistical power. Second, TTT measures an exponential amplification process; its relationship with initial seed concentration and disease progression might not be perfectly linear. Finally, substantial inter-individual variability in seeding kinetics might become more apparent—and potentially confounding—in larger sample sizes.

Several limitations should be acknowledged. First, this study was based on observational data from the PPMI cohort and cannot establish causality. Second, the study was conducted using the PPMI cohort, which consists predominantly of White participants and may not fully reflect the PD population encountered in routine clinical practice. Therefore, validation in independent longitudinal cohorts and more diverse populations will be necessary to establish the generalizability and reproducibility of our findings.

## Conclusion

αSyn-SAA positivity was associated with motor deterioration and phenoconversion during the prodromal phase. These findings suggest that the relationship between pathological α-synuclein and clinical progression is stage-dependent and may be strongest during the prodromal period, when disease progression remains more closely linked to α-synuclein pathology. Beyond its established roles as a diagnostic and risk biomarker in PD and other synucleinopathies, αSyn-SAA may therefore serve as a biomarker of disease progression during the prodromal phase and as a valuable tool for the design of future α-synuclein-targeted therapeutic trials.

## Supporting information

Supplementary results 1

Supplementary results 2

## Data Availability

Data used in the preparation of this article were obtained on 2026-07-29 from the Parkinson’s Progression Markers Initiative (PPMI) database (https://www.ppmi-info.org/access-data-specimens/download-data), RRID:SCR_006431. For up-to-date information on the study, visit https://www.ppmi-info.org/. Data are available to qualified researchers through the PPMI database, subject to the PPMI data access requirements.

## Acknowledgement

PPMI – a public-private partnership – is funded by the Michael J. Fox Foundation for Parkinson’s Research and funding partners, including AbbVie, Alamar Biosciences, Aligning Science Across Parkinson’s (ASAP), Arrowhead Pharma, Arvinas, AskBio, BIAL, BioArctic, Biohaven, BlueRock Therapeutics, Bristol Myers Squibb, Calico Labs, Capsida Biotherapeutics, Critical Path Institute, DaCapo Brainscience, Denali, Edmond J. Safra Foundation, Eli Lilly, Gain Therapeutics, GE Healthcare, Genentech, GSK, Insitro, Johnson & Johnson Innovative Medicine, Lundbeck, Merck, Neumora, Neuron23, Novartis, Olink, Regeneron, Roche, Sanofi, Tenvie, UCB, Vanqua Bio, Voyager Therapeutics, and The Weston Family Foundation.

