## Supplementary results 1 for "α-synuclein seed amplification assay positivity is associated with motor progression in the prodromal stage independent of phenoconversion"

**Supplementary result 1:**

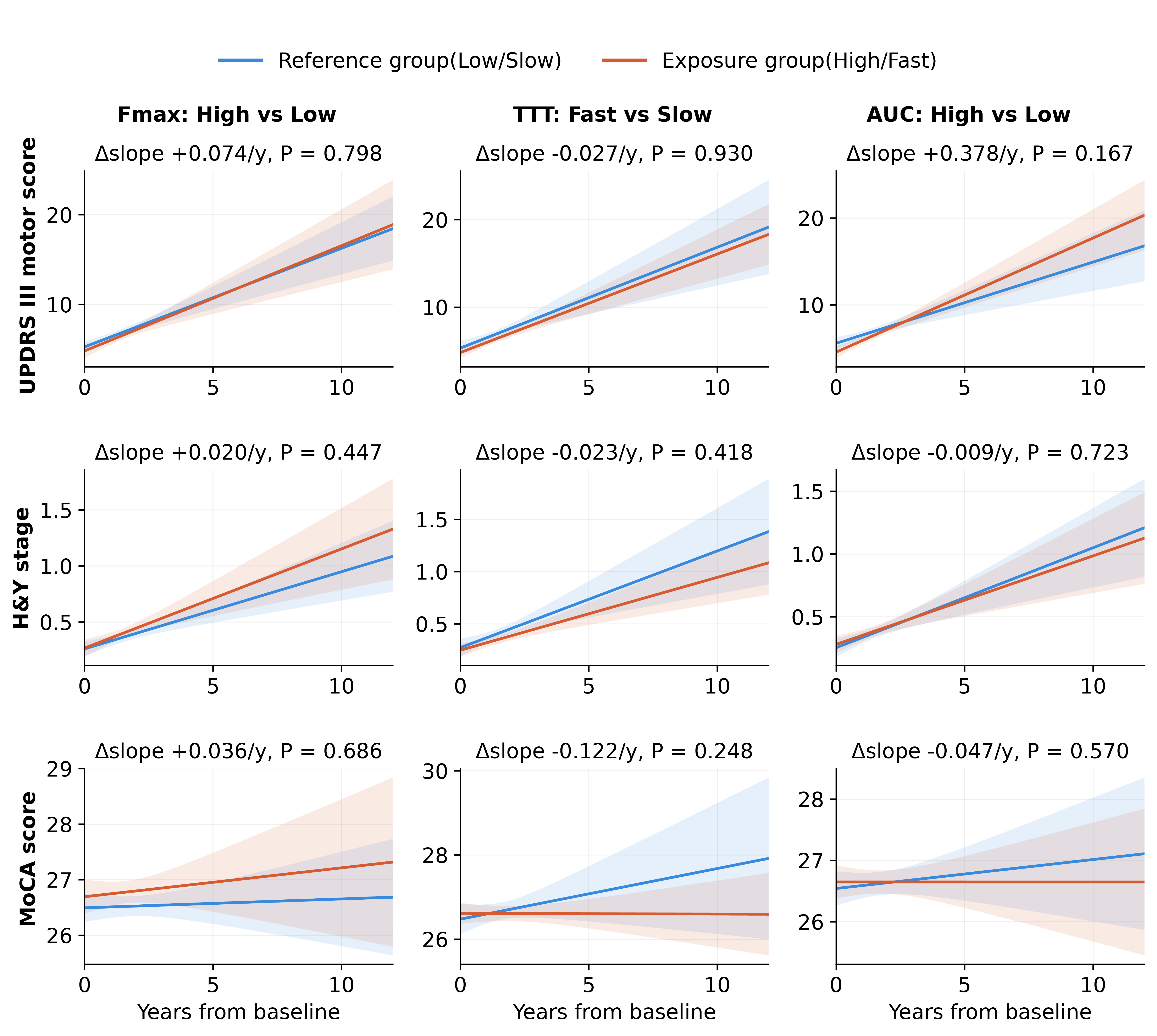

**Figure S1. Longitudinal progression of clinical outcomes according to baseline SAA kinetic parameters.** Model-estimated trajectories for MDS-UPDRS Part III motor score, H&Y stage, and MoCA score are shown for high/fast (orange) and low/slow (blue) groups based on baseline Fmax, TTT, and AUC among SAA-positive participants. Shaded areas represent 95% CIs. Δslope represents the estimated difference in annual rate of change between high/fast and low/slow groups; P values test the difference in slopes.
