## Supplementary results 2 for "α-synuclein seed amplification assay positivity is associated with motor progression in the prodromal stage independent of phenoconversion"

**Supplementary result 2:**

Compared with Coughlin et al. (2025),^1^ the updated analysis included a substantially larger prodromal cohort (1,796 vs. 96 participants), more phenoconversion events (181 vs. 23), and longer follow-up (12 vs. 9.2 years; Table S1). SAA positivity remained significantly associated with phenoconversion (P<0.001), with an adjusted HR of 5.72 (95% CI 3.78–8.68). However, the previously significant difference between slow and fast seeders (P=0.03) was no longer statistically significant in the updated analysis (adjusted HR 0.73, 95% CI 0.51–1.03; P=0.074).

Figure S1 shows the model-estimated longitudinal trajectories of UPDRS part III motor score, H&Y stage, and MoCA score according to baseline Fmax, TTT, and AUC kinetic groups. Analyses were adjusted for age, sex, family history, phenoconversion status, and the corresponding baseline clinical score. The MoCA model was additionally adjusted for education years and APOE ε4. No significant differences in longitudinal slopes were observed between groups for any of the clinical outcomes (all P > 0.05).

**Table S1. Comparison of key findings between the original and updated analyses**

| **Characteristic** | **Coughlin et al., 2025** | **Updated results** |
| --- | --- | --- |
| Prodromal cohort, total N | 96 | 1796 |
| αSyn-SAA positive, n (%) | 54 (56.2%) | 1040 (57.9%) |
| αSyn-SAA negative, n | 42 | 756 |
| Phenoconversion events, total, n | 23 | 181 |
| Maximum follow-up, y | 9.2 | 12 |
| SAA+ vs SAA-, log-rank p | 0.001 | P < .001 |
| Adjusted HR, SAA+ vs SAA-, (95% CI) ^†^ | Not reported | 5.72 (3.78–8.68) |
| Fast seeders, n (events) | 30 (16) | 496 (81) |
| Slow seeders, n (events) | 24 (5) | 495 (61) |
| Adjusted HR, Fast vs Slow (95% CI) ^†^ | Not reported | 0.73 (0.51–1.03) |
| Fast vs Slow, p-value ^†^ | 0.03 | 0.074 |

*All available follow-up was used in the survival and Cox analyses.

†Age- and sex-adjusted Cox proportional hazards model; the reference group is αSyn-SAA-negative or Slow seeders, respectively.

1. Coughlin DG, Shifflett B, Farris CM, et al. alpha-Synuclein Seed Amplification Assay Amplification Parameters and the Risk of Progression in Prodromal Parkinson Disease. *Neurology*. Mar 11 2025;104(5):e210279. doi:10.1212/WNL.0000000000210279

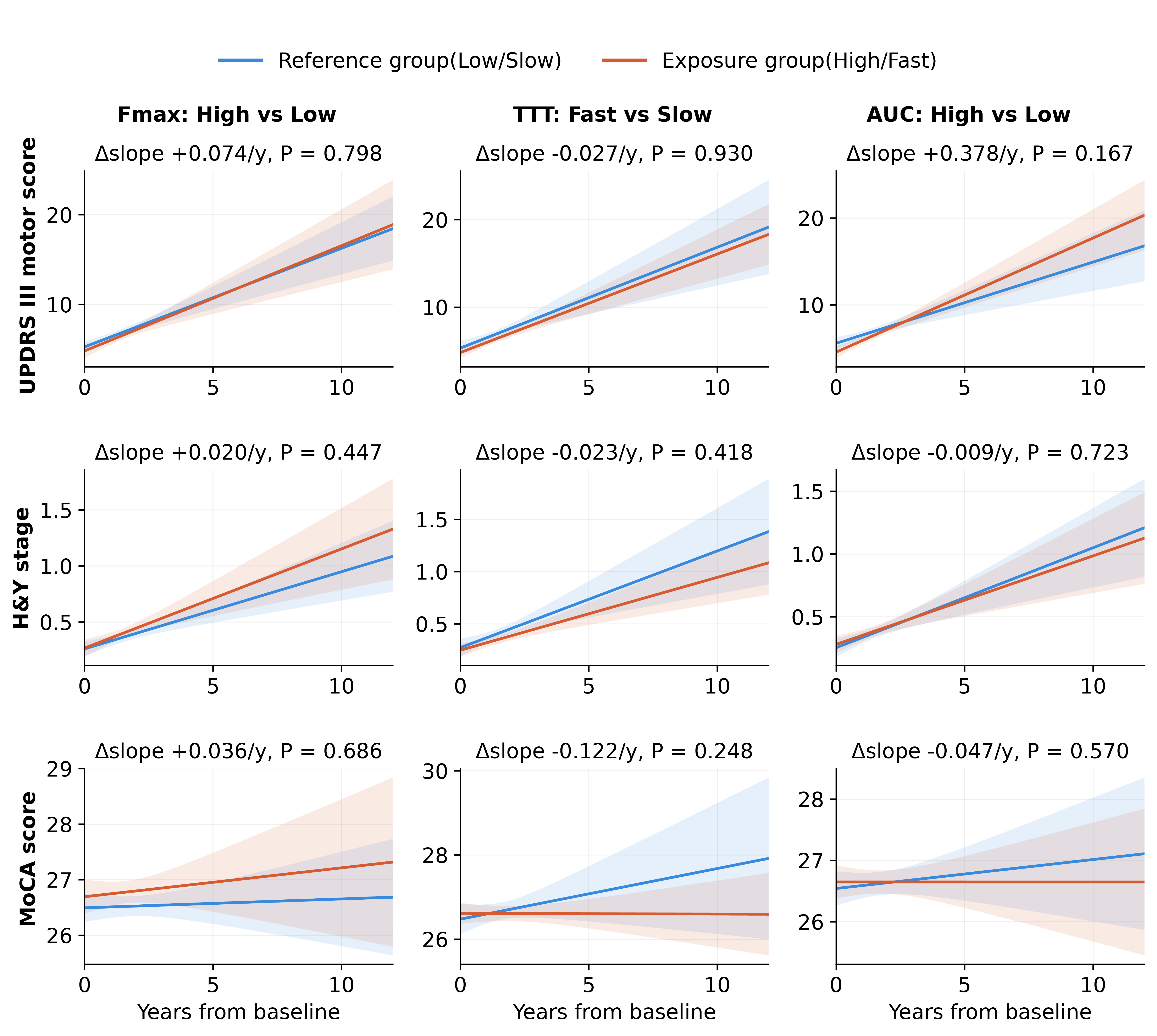

**Figure S1. Longitudinal progression of clinical outcomes according to baseline SAA kinetic parameters.** Model-estimated trajectories for MDS-UPDRS Part III motor score, H&Y stage, and MoCA score are shown for high/fast (orange) and low/slow (blue) groups based on baseline Fmax, TTT, and AUC among SAA-positive participants. Shaded areas represent 95% CIs. Δslope represents the estimated difference in annual rate of change between high/fast and low/slow groups; P values test the difference in slopes.
